# Comparison of Omicron Breakthrough Infection Versus Monovalent SARS-CoV-2 Intramuscular Booster Reveals Differences in Mucosal and Systemic Humoral Immunity

**DOI:** 10.1101/2023.09.22.23295541

**Authors:** Sabryna Nantel, Salma Sheikh-Mohamed, Gary Y. C. Chao, Alexandra Kurtesi, Queenie Hu, Heidi Wood, Karen Colwill, Zhijie Li, Ying Liu, Laurie Seifried, Benoîte Bourdin, Allison McGeer, William R. Hardy, Olga L. Rojas, Mario A. Ostrowski, Mark A. Brockman, Ciriaco A. Piccirillo, Caroline Quach, James M. Rini, Anne-Claude Gingras, Hélène Decaluwe, Jennifer L. Gommerman

## Abstract

Our understanding of the quality of cellular and humoral immunity conferred by COVID-19 vaccination alone versus vaccination plus SARS-CoV-2 breakthrough (BT) infection remains incomplete. While the current (2023) SARS-CoV-2 immune landscape of Canadians is complex, in late 2021 most Canadians had either just received a third dose of COVID-19 vaccine, or had received their two dose primary series and then experienced an Omicron BT. Herein we took advantage of this coincident timing to contrast cellular and humoral immunity conferred by three doses of vaccine versus two doses plus BT. Our results show that mild BT infection induces cell-mediated immune responses to variants comparable to an intramuscular vaccine booster dose. In contrast, BT subjects had higher salivary IgG and IgA levels against the Omicron Spike and enhanced reactivity to the ancestral Spike for the IgA isotype, which also reacted with SARS-CoV-1. Serum neutralizing antibody levels against the ancestral strain and the variants were also higher after BT infection. Our results support the need for mucosal vaccines to emulate the enhanced mucosal and humoral immunity induced by Omicron without exposing individuals to the risks associated with SARS-CoV-2 infection.

**ONE SENTENCE SUMMARY:** Omicron breakthrough elicits cross-reactive systemic and mucosal immune responses in fully vaccinated adults.

**Graphical Abstract:** 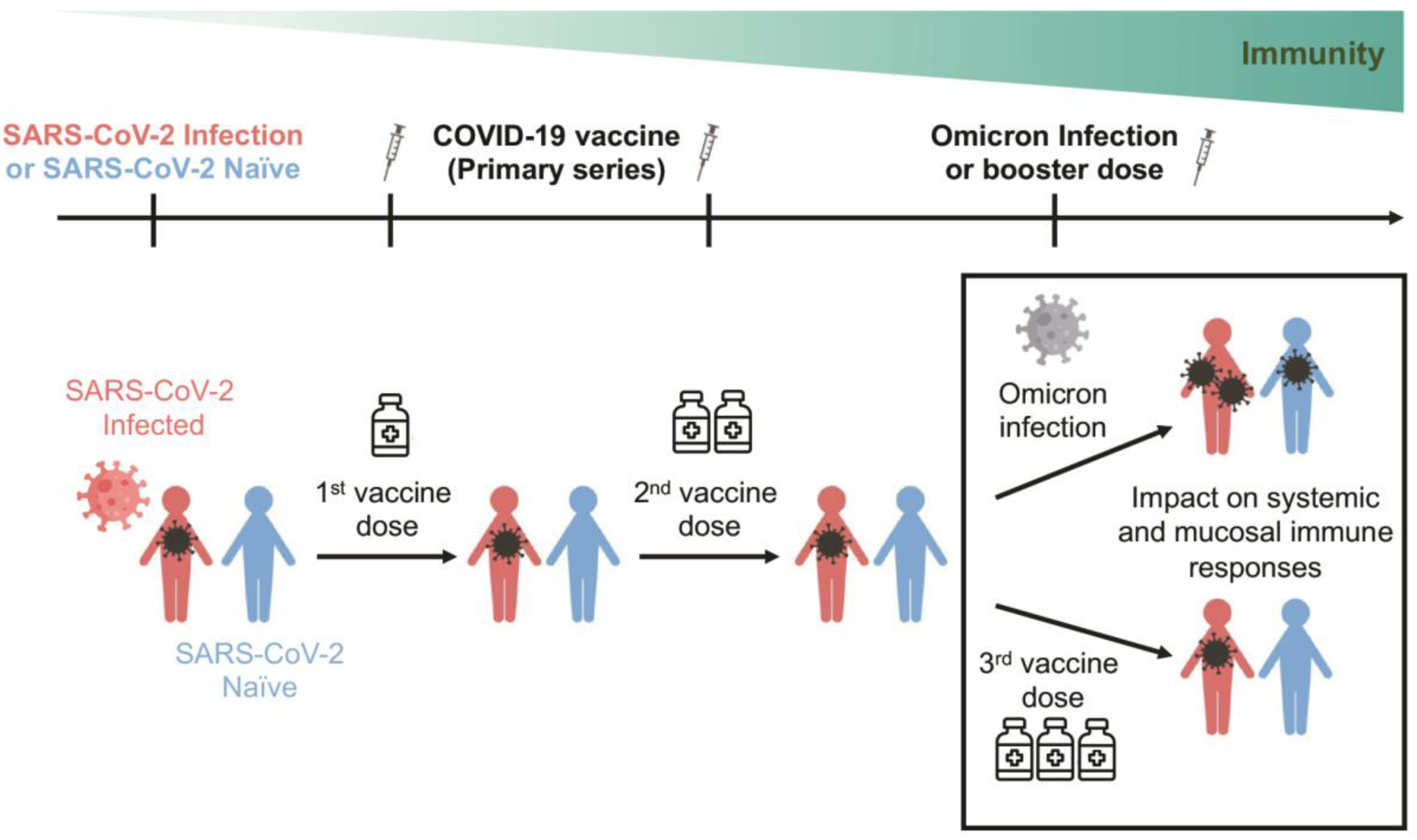

## INTRODUCTION

SARS-CoV-2 vaccines provide robust protection against severe illness and death, making them crucial aids in managing COVID-19. Multiple studies have assessed both mucosal and systemic immune responses to SARS-CoV-2 vaccination and found that vaccination results in a strong plasmablast response, with neutralizing antibodies (nAb) serving as a reasonable correlate of protection.^1^ However, as the initial plasmablast response subsides, circulating antibody (Ab) levels naturally decline.^2^ Furthermore, as increasingly divergent variants appear, transmission dynamics of SARS-CoV-2 change.^3,4^ Increased transmissibility of the Omicron variant which emerged in December 2021, coupled with declines in vaccine-induced circulating Ab titers, resulted in a skyrocketing of breakthrough (BT) cases.^5^

We previously showed that a single dose of COVID-19 mRNA vaccine induced a detectable but low amount of salivary Spike and RBD-specific IgA that did not persist in most people, and that salivary anti-Spike and anti-RBD-specific IgA highly correlated with salivary secretory IgA.^6^ In contrast, a combination of prior SARS-CoV-2 infection followed by vaccination, refered to as “hybrid immunity”, induced more robust salivary Spike and RBD-specific IgA.^7,8^ The impact of BT infection on the pre-existing immune response, and particularly mucosal immunity, as well as the breadth of this response against other variants or Betacoronaviruses is still unclear.

For most people living in Canada, third doses of COVID-19 vaccines were rolled out in December 2021, at the same time that Omicron emerged. We took advantage of this coincident timing to assess humoral and cell-mediated immune responses in two cohorts of participants who received two doses of vaccine compared to two doses of vaccine and an Omicron BT or three doses of vaccine (primary series followed by booster dose). One cohort of participants were infection-naïve, and the second cohort were infected in the pre-vaccine era (2020).^9^ We assessed serum for the presence of binding and neutralizing Ab, saliva for the presence of binding Ab and peripheral blood mononuclear cells (PBMCs) for interferon-gamma (IFN-γ) production by T cells in response to different SARS-CoV-2 variants. Our results demonstrate that an Omicron BT infection induces higher and more broadly neutralizing antibody titers compared to an intramuscular vaccine boost only in previously uninfected subjects. We also found that a BT induced salivary anti-Spike IgA against SARS-CoV-2 and its Omicron variant, but also cross-reactive IgA against SARS-CoV-1, a distinct Betacoronavirus. Finally, Omicron BT strongly boosted T-cell responses, even in individuals with prior hybrid immunity. Overall, our results confirm the impact and benefits of combining multiple routes of antigenic stimulation to support strong systemic and mucosal immune responses.

## RESULTS

### Study design

The data in this study was collected from two different cohorts **(Fig. 1)**. The first cohort (CoVaRR-Net or CVR Cohort, **Supplemental Table 1**) consist of previously naïve individuals who subsequently received two (2x Vax) or three (3x Vax) doses of vaccine, or two vaccine doses followed by an Omicron BT infection (2x Vax + BT) **(Fig. 1A)**. The vaccines administered in the CVR cohort were a mix of mRNA vaccines (Pfizer-BioNTech’s BNT162b2 and Moderna’s mRNA1273), as well as a viral vector vaccine (AstraZeneca-Oxford’s ChAdOx1), reflecting the real-world composition of Canadian vaccinees. The second cohort (RECOVER Cohort, **Supplemental Table 2**) is composed of health care workers who experienced a SARS-CoV-2 infection prior to vaccination and subsequently received either two (Inf. + 2x Vax) or three vaccine doses (Inf. + 3x Vax) or two doses followed by an Omicron BT infection (Inf. + 2x Vax + BT) **(Fig. 1B)**.^9^ All participants in the RECOVER cohort were administered Pfizer-BioNTech’s BNT162b2 mRNA vaccine. Serum and peripheral blood mononuclear cells (PBMCs) were collected for both cohorts, while saliva was available for the CVR cohort only **(Fig. 1B-C)**. Serum samples were used to assess SARS-CoV-2-specific binding and neutralizing Ab titers, while PBMCs were used to assess SARS-CoV-2-specific functional T-cell responses measured through Enzyme-Linked ImmunoSpot Assay (ELISpot) for the detection of IFN-γ secreting cells **(Fig. 1C)**. Saliva was used to measure mucosal Spike and RBD-binding Abs **(Fig. 1D)**.

**Figure 1.**
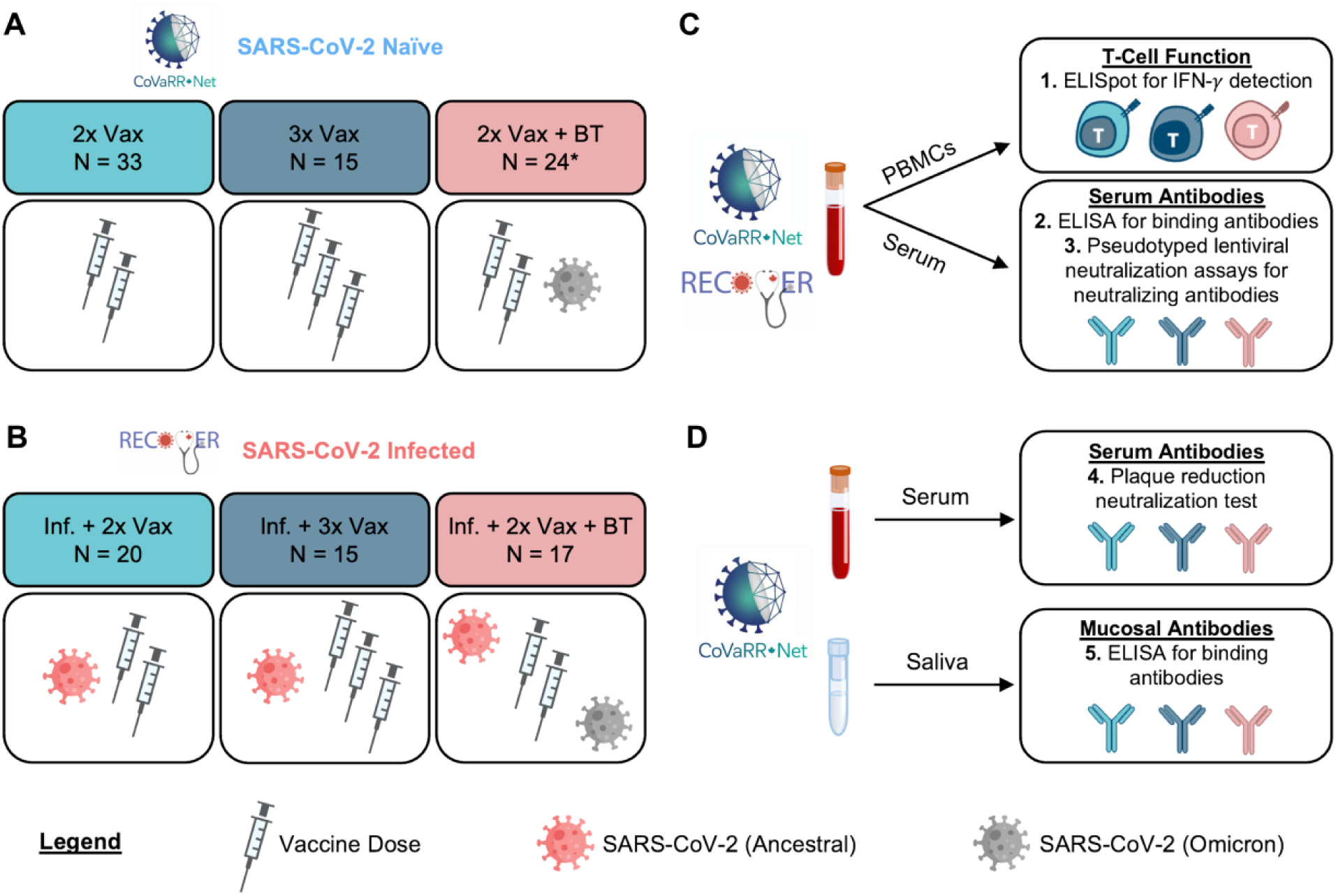
Cohorts, Sampling and Experimental Set-Up. Blood samples were drawn from three groups in two distinct cohorts. Omicron naïve individuals who received two or three vaccine doses (Vax, light and dark blue) and individuals vaccinated twice who had an Omicron breakthrough (BT, red) at the time of BA.1 dominance in cohorts **(A)** without prior SARS-CoV-2 infection (CoVaRR-Net/CVR cohort) and **(B)** with SARS-CoV-2 infection prior to vaccination (RECOVER cohort). The cohort without prior exposure to SARS-CoV-2 was vaccinated with BNT162b2, mRNA1273 and/or ChAdOx1 vaccines, while the previously infected cohort was vaccinated with BNT162b2 only. Plasma and PBMCs were collected after vaccination or infection. The CVR cohort also had concurrent saliva collection. The demographic composition of each cohort is further detailed in **Supplemental Tables 1 and 2**. **(C)** Three assays were performed for both cohorts : 1. Interferon-*γ* producing T-cells in response to SARS-CoV-2 peptides were quantified using ELISpot assays. 2. Serum antibody levels were measured using an ELISA-based detection method. 3. Serum neutralizing capacity was assessed using pseudotyped lentiviral neutralization assays. **(D)** Two additional assays were performed for the CoVaRR-Net cohort only : 4. Serum neutralizing capacity was independently-confirmed using plaque reduction neutralization test. 5. Mucosal antibody levels in saliva were measured with a biotinylated ELISA-based detection method. *This group includes salivary-analysis-only samples.

### Omicron breakthrough infection enhances T-cell responses in infection-naïve individuals and individuals with prior hybrid immunity who had received two doses of vaccine

We assessed functional T-cell immunity in response to Spike peptides from the ancestral Wuhan strain of SARS-CoV-2, as well as subsequent variants of concern (Alpha, Beta, Gamma, Delta and Omicron BA.1). All participants who received three doses of vaccines or developed a BT infection after their primary series displayed Spike-specific T-cell responses above the positive threshold value, irrespective of the cohort from which they were derived. For all six strains of SARS-CoV-2 Spike assessed, an Omicron BT infection significantly increased the number of IFN-γ secreting cells compared to individuals who were only vaccinated twice (**Fig. 2 and Fig. S1**). The impact of BT infection on the breadth of T-cell immune response was irrespective of previous viral exposure, as individuals initially infected with the ancestral Wuhan strain also exhibited a two-fold increase in IFN-γ secreting cells following BT infection compared to individuals who received two vaccine doses (**Fig. 2)**. Altogether, a BT infection or a third vaccine dose induced comparable functional T-cell responses in both cohorts. Notably, a BT infection enhanced T-cell responses even in individuals who were infected prior to their primary series and were therefore already hybrid immune.

**Figure 2.**
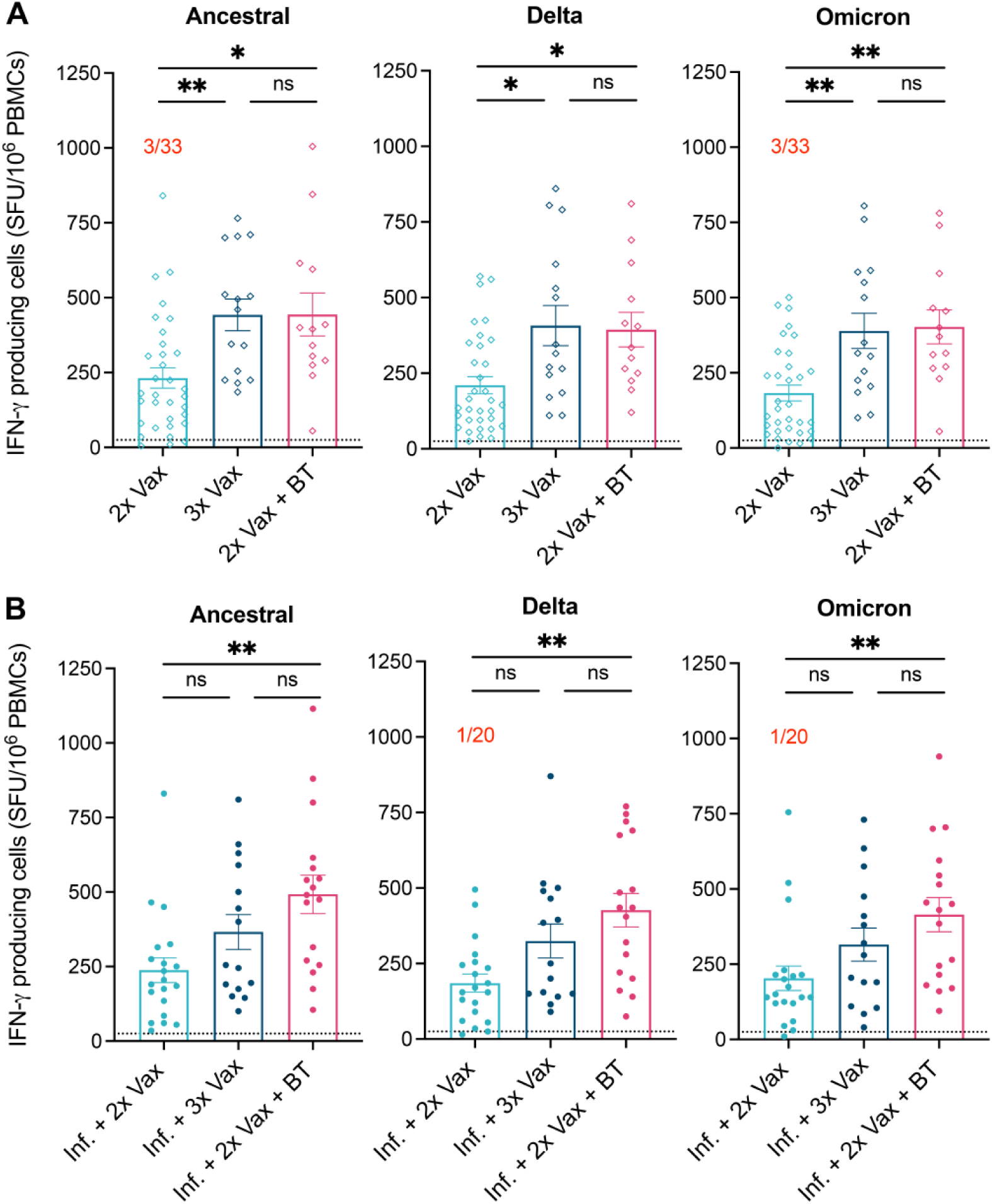
Omicron breakthrough infection boosts T-cell responses to SARS-CoV-2 and its variants in vaccinated and hybrid immune individuals who had received two doses of vaccine. Samples were collected from **(A)** previously naïve individuals (empty diamonds) and **(B)** individuals infected with the ancestral SARS-CoV-2 (full circles). PBMCs were analyzed after two (n=33, light blue), three (n=15, dark blue) or two vaccine doses followed by breakthrough (BT) infection (n=13, red) for the cohort naïve to SARS-CoV-2. PBMCs were analyzed after two (n=20, light blue), three (n=15, dark blue) or two vaccine doses of BNT162b2 followed by an Omicron BT infection (n=17, red) for the cohort infected (Inf.) prior to vaccination. T-cell responses were assessed by ELISpot after SARS-CoV-2 Spike peptide stimulation with the Ancestral (Wuhan-1 Like, left panels) or variant (Delta and Omicron, middle and right panels respectively) strains. Results are expressed in number of IFN-γ producing cells per million PBMCs. Dotted line indicates the positive threshold value. The number of participants with responses under the positive cut-off value are indicated in red for each group when it applies. Error bars indicate mean ± SEM. Statistical significance was established as : ns (not significant) P >.05, *P <.05, **P <.01.

### Omicron breakthrough infection enhances circulating anti-Spike and anti-RBD antibody levels in vaccinated individuals without prior infection

We next measured serum antibody levels in infection-naïve participants (**Fig. 3A-B)** and hybrid-immune subjects (**Fig. 3C-D**). Similar to what was observed for T-cell responses, an Omicron BT boosted IgG and IgA levels beyond what was observed with two doses of vaccine, even in individuals who were infected before receiving their primary vaccine series. In the infection-naïve cohort, IgG and IgA antibody levels against SARS-CoV-2 Ancestral Spike and RBD proteins were significantly higher in participants who received two doses of vaccine followed by BT than those who received two or three doses of vaccine (**Fig. 3A-B**). In contrast, in the hybrid-immune cohort, although BT infection elicited higher serum antibody levels compared to the ones observed after two vaccine doses, a third vaccine dose led to antibody levels comparable to BT infection (**Fig. 3C-D**).

**Figure 3.**
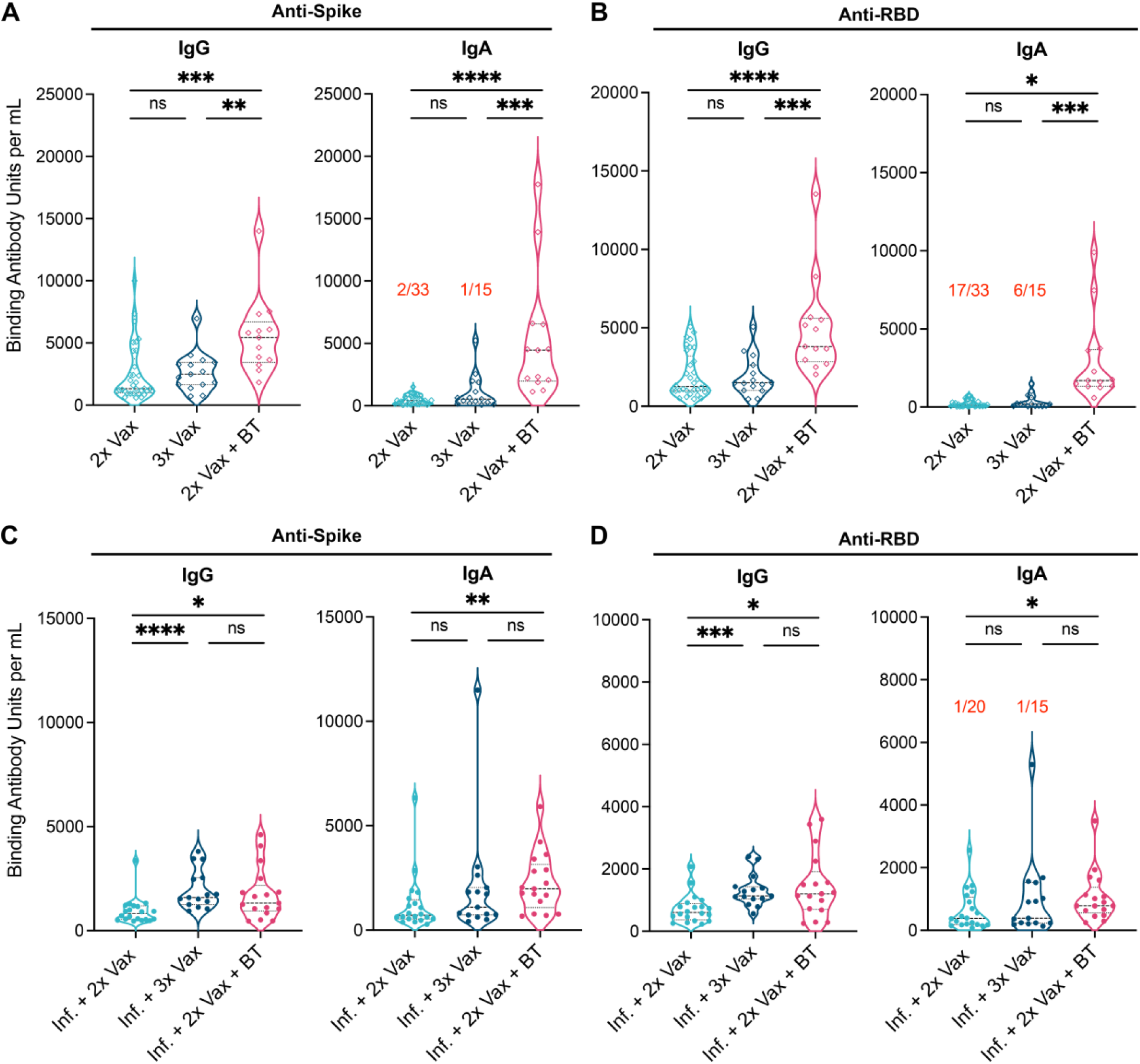
Omicron BT infection enhances humoral immune responses in vaccinated individuals who were previously naïve to the infection. Anti-Spike **(A, C)** and Anti-RBD **(B, D)** IgG (left panels) and IgA (right panels) levels were measured in the serum of previously naïve (empty diamonds, A and B) and previously infected individuals (full circles, C and D). Serum were analyzed after two (n=33, light blue), three (n=15, dark blue) or two vaccine doses followed by breakthrough (BT) infection (n=13, red) for the cohort naïve to SARS-CoV-2. Serum were analyzed after two (n=20, light blue), three (n=15, dark blue) or two vaccine doses of BNT162b2 mRNA vaccine followed by an Omicron BT infection (n=17, red) for the cohort infected (Inf.) prior to vaccination. The number of participants with responses under the positive cut-off value are indicated in red for each group when it applies. Statistical significance was established as : ns (not significant) P >.05, *P <.05, **P <.01, ***P<.001, ****P<.0001.

In addition to binding Ab levels, neutralizing antibody titers were assessed using Spike-pseudotyped lentiviral assays against multiple Omicron subvariants (BA.1, BA.2 and BA.5) as well as the Ancestral, Beta and Delta strains in both cohorts (**Fig. 4**). In infection-naïve vaccinated participants, BT infection resulted in significantly increased neutralizing capacity against all variants compared to two doses of vaccine, and to all variants except Omicron BA.2 compared to three doses of vaccine (**Fig. 4A**). In contrast, in hybrid immune individuals, although BT infection also elicited higher neutralizing antibody titers compared to two doses of vaccine, BT infection did not result in enhanced neutralizing capacity compared to three doses of vaccine (**Fig. 4B**). The results of this pseudotyped lentiviral neutralization assay were further supported by an independently-conducted plaque reduction neutralization test (**Fig. S2**). Taken together, in participants who have received two doses of vaccine, BT infection induces higher levels of binding and neutralizing Ab to Spike and RBD than a third dose of vaccine. However, this is only the case for participants who were never infected prior to their vaccination series.

**Figure 4.**
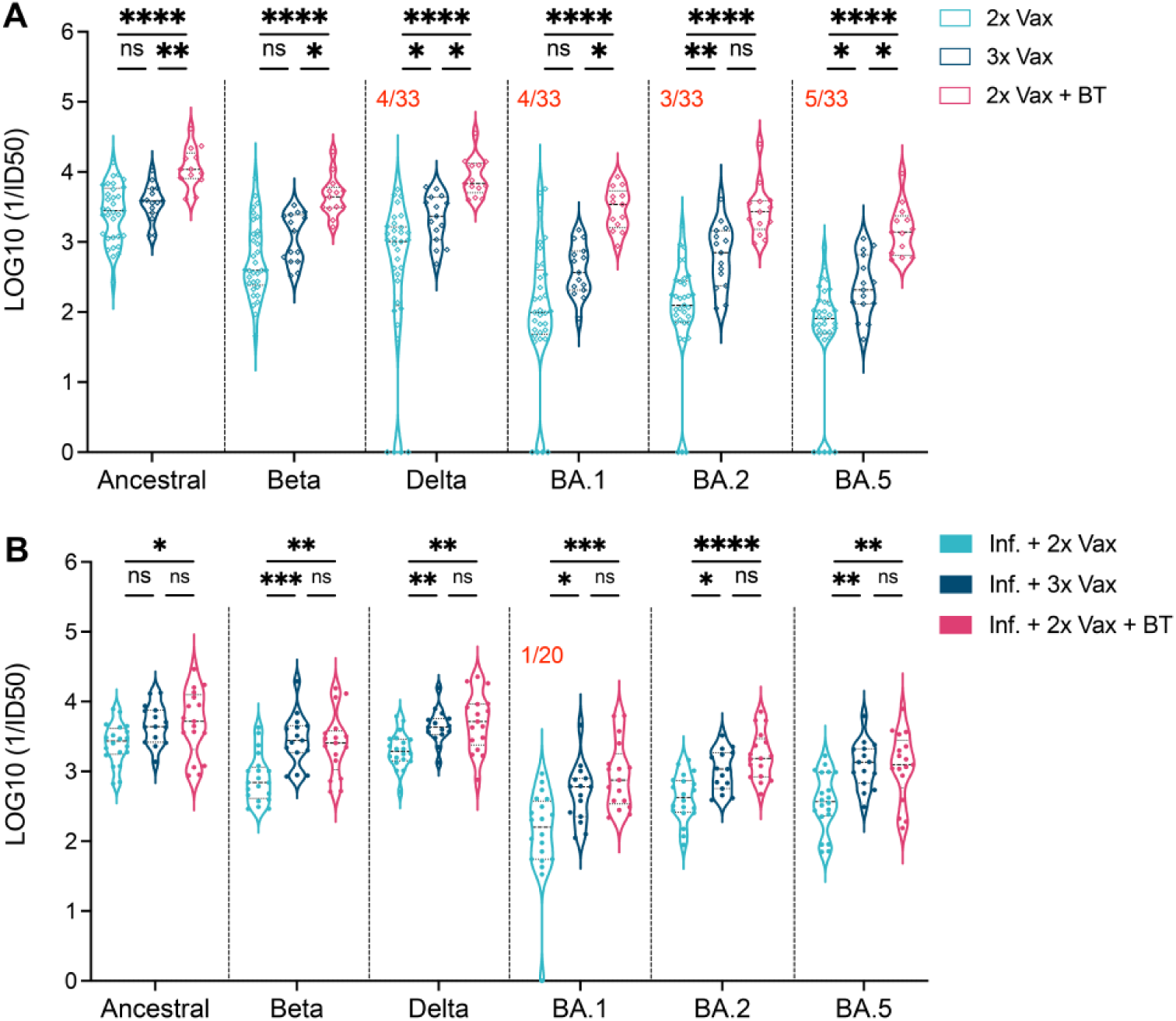
Omicron breakthrough infection increases neutralizing capacity against several variants in vaccinated individuals who were previously naïve to the infection. Spike-pseudotyped lentivirus neutralization assays were conducted to establish the serum dilution that neutralized the infectivity of Spike-pseudotyped lentivirus particles of different variants (Ancestral, Beta, Delta, BA.1, BA.2 and BA.5). **(A)** Serums were analyzed after two (n=33, light blue), three (n=15, dark blue) and two vaccine doses followed by breakthrough (BT) infection (n=13, red) in individuals who were naïve to SARS-CoV-2 (empty diamonds). **(B)** Serums were analyzed after two (n=20, light blue), three (n=15, dark blue) and two vaccine doses of BNT162b2 vaccine followed by an Omicron breakthrough infection (n=17, red) in individuals infected (Inf.) prior to vaccination (full circles). The number of participants without detectable neutralizing antibodies against specific variants are indicated in red for each group when it applies. Statistical significance was established as : ns (not significant) P >.05, *P <.05, **P <.01, ***P<.001, ****P<.0001.

### An Omicron breakthrough infection induces cross-reactive salivary IgA

We next measured binding IgG and IgA levels against SARS-CoV-2 Spike and RBD in the saliva of participants who were naïve to the infection prior to vaccination and were subsequently boosted with a third vaccine dose or experienced an Omicron BT (**Fig. 5**). We normalized antigen-specific IgA and IgG levels to total albumin protein in the saliva of each subject which, unlike total IgA, was similar between groups (**Fig. S3**). IgG and IgA levels to Omicron Spike and Omicron RBD mostly undetectable in participants who experienced an Omicron BT (**Fig. 5**). While Omicron BT infection did not impact salivary IgG against the ancestral Spike and RBD beyond what was observed in participants who received three doses of vaccine (**Fig. 5A**), IgA against both ancestral Spike and RBD were significantly elevated in BT participants (**Fig. 5B**). Unexpectedly, SARS-CoV-2 BT infection also induced IgA against SARS-CoV-1 Spike and RBD (**Fig. 5B**). Overall, two doses of vaccine followed by BT infection induced higher and broader IgA levels in saliva compared to vaccination alone.

**Figure 5.**
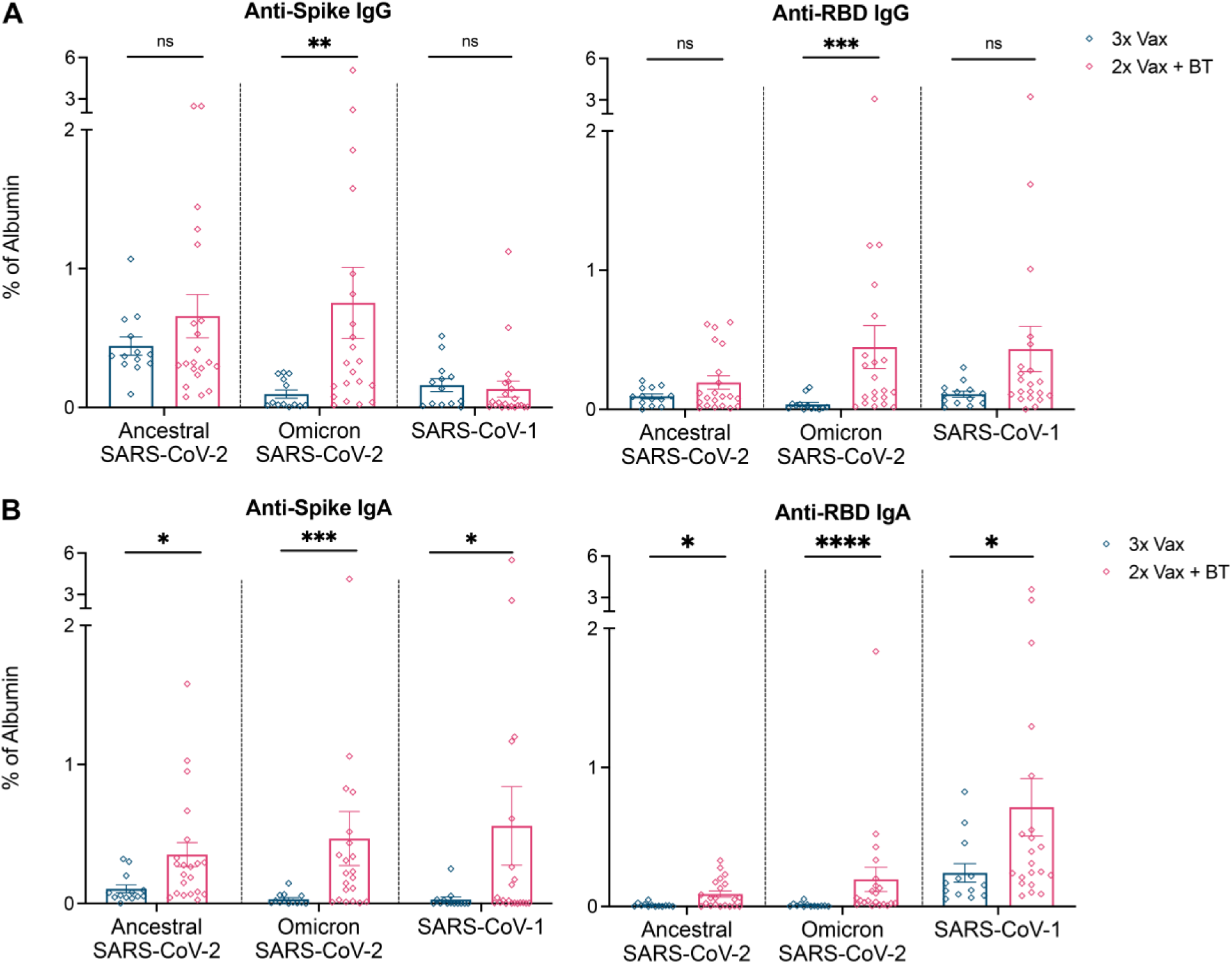
Omicron breakthrough infection induces a stronger and more diverse mucosal immune response than an intramuscular vaccine booster. Anti-Spike IgG (A, left panel) and IgA (B, left panel), as well as anti-RBD IgG (A, right panel) and IgA (B, right panel) antibody responses to the ancestral SARS-CoV-2, SARS-CoV-2 Omicron variant and SARS-CoV-1 were analyzed after three (n=14, dark blue) or two vaccine doses followed by an Omicron breakthrough (BT) infection (n=21, red) in individuals previously naïve to SARS-CoV-2 (empty diamonds). Results are expressed as a percentage normalized to albumin detected in saliva. Error bars indicate mean ± SEM. Statistical significance was established as : ns (not significant) P >.05, **P <.01, ***P<0.001, ****P<0.0001.

## DISCUSSION

In this study, we took advantage of the synchronous timing of the beginning of the booster vaccination campaign and the emergence of Omicron in Canada, both of which occurred in December 2021, to determine if two doses of vaccine followed by BT infection conferred comparable immunity to three doses of vaccine. While the breath and magnitude of T-cell responses were comparable in these two groups, humoral immunity was enhanced in participants who had two doses of vaccine followed by BT. However, this enhancement was only observed in the absence of SARS-CoV-2 infection prior to vaccination. Therefore, the systemic humoral immunity advantage of a localized intranasal infection was lost in participants who had already experienced an infection prior to vaccination and thus, had already developed an hybrid immune response. Notably, BT infection conferred higher levels of salivary IgA against non-Omicron forms of Spike, including the ancestral Wuhan-like Spike and even SARS-CoV-1 Spike, compared to an intramuscular vaccine boost. immunity.

Our results also revealed that an Omicron BT infection was able to enhance T-cell responses to multiple variants even in individuals with prior hybrid immunity generated by an initial SARS-CoV-2 infection and a primary immunization series of two doses. This suggests that individuals with hybrid immunity can still benefit from mucosal boosting to sustain functional memory T-cell responses to SARS-CoV-2 and its multiple variants. This is particularly important for individuals with prior asymptomatic infections, as we have shown that these asymptomatic infections do not prime the immune system as efficiently as symptomatic ones.^10^ Moreover, the demonstration that BT infection potently enhanced T-cell responses irrespectively of previous exposure history supports the need for development of mucosal vaccine boosters to sustain the breath and longevity of the memory T-cell pool to SARS-CoV-2 variants. Indeed, when compared to other studies, our results suggest that an Omicron BT behaves as an intranasal vaccine boost, both improving the levels of SARS-CoV-2 specific T cells and the broadness of the antibody response, with detection of antibodies against SARS-CoV-1.^11^

Similar to other studies, we found that BT infection confered a rise in cellular and humoral immunity against virus strains that participants had not experienced, such as Delta, beyond what is observed with a primary vaccine series.^12,13^ In the case of T cell recall to other variant strains, it has been shown that CD4^+^ and CD8^+^ T cells induced by previous infection or vaccination could cross-recognize other variants such as the Omicron B.1.1.529 while maintaining similar phenotype and functionality.^14^ In the case of the humoral immune response, it has been postulated that such an enhanced memory response with broadened reactivity could be due to the back-boosting of memory B cells that react to conserved Spike epitopes.^12,13^

We also observed that, in previously uninfected participants, Omicron BT infection augmented the nAb response to most variants tested compared to double or triple-vaccinated participants. However, in participants infected with SARS-CoV-2 prior to vaccination, BT infection did not elevate nAb levels to SARS-CoV-2 variants beyond what was observed for individuals infected and vaccinated with three doses. From this, we can conclude that infection followed by a primary series and a booster dose confers a maximal nAb response against the variants tested and is not improved by further exposure to Omicron. These data agree with other reports showing that multiple antigenic exposures confer a “ceiling” of the nAb response.^12,13^ Since we did not observe an added benefit of Omicron BT on the nAb titers in participants who were exposed to ancestral SARS-CoV-2 prior to vaccination, we can conclude that the generation of the nAb “ceiling” is not unique to Omicron exposure. Rather, our results suggest that one mucosal exposure is required to generate maximal nAb levels against different SARS-CoV-2 variants.

Considering the high transmission rates and infection numbers during the Omicron wave, it is clear that vaccine-induced nAb do not necessarily provide protective immunity sufficient to prevent infection against this divergent variant.^15,16^ Indeed, Tang *et al.* found that vaccination induced only low levels of mucosal antibodies in the upper respiratory tract and that neutralizing antibodies collected from bronchiolar lavage fluid were almost completely ineffective against the Omicron BA.1 variant.^8^ Furthermore, it appears that Spike and RBD-specific salivary secretory IgA (SIgA) are weakly induced by vaccination alone, but require mucosal exposure to SARS-CoV-2 for higher titres.^7,17^ This highlights the importance of vaccination strategies that take into account the induction of nAb in the upper respiratory tract, the site of viral encounter. Indeed, it has been shown that high salivary and nasal Spike-specific IgA is associated with protection against BT infection.^18,19^ We show here that Omicron BT infection induced salivary Ab that not only bind the Omicron Spike protein, but also induced IgA that bind to the ancestral and SARS-CoV-1 Spike protein. Due to the absence of saliva sampling in the cohort infected prior to their vaccination series, we do not know whether hybrid immunity induced by this initial infection induces long-lasting salivary IgA of similar breadth. Moreover, we cannot make conclusions on the impact of the order of virus exposure, (ie, before or after vaccination), on salivary antibody production.

One potential weakness of this study is that, although vaccine combinations were carefully matched between groups for each cohort, these cohorts included participants who were administered different vaccines (**Supplemental Tables 1 and 2**). While the infection-naïve cohort was collected in Toronto and included BNT162b2, mRNA-1273 and ChAdOx1-S recipients, the cohort with infection prior to vaccination was independently collected in Montreal and only included BNT162b2 recipients. However, these cohorts reflect the heterogeneous immune landscape of the Canadian population.^20^ Despite these differences, results from these cohorts suggest that while repeated intramuscular (monovalent) vaccination does not strikingly improve the broadness of the response, mucosal exposure induces more potent immune responses compared to vaccinated-only individuals. Another weakness of our study is that the relative durability of B and T-cell memory following BT compared to booster dose is not known, as serum samples were collected approximately six weeks following either an Omicron BT or a third vaccine dose. Such studies, while valuable, are increasingly challenging to conduct due to our inability to discern whether asymptomatic infections have occurred in hybrid immune populations, and to control for multiple BT infections.

In conclusion, we provide evidence that an Omicron BT induces higher and broader nAb titers compared to three doses of COVID-19 vaccine. We also showed that an Omicron BT supports the persistence of functional memory T cells which can efficiently cross-recognize multiple variants of SARS-CoV-2. Lastly, we find an added benefit of mucosal exposure over intramuscular vaccination in the form of salivary IgA detected in the oral cavity following infection and capable of recognizing divergent Spike proteins. Therefore, our results strongly support the development of intranasal vaccination approaches to enhance and broaden long-lasting immune responses to SARS-CoV-2 and its multiple variants.

## METHODS

### Study Approvals

The University of Toronto (UofT) Research Ethics Board (REB) granted approval for recruitment of participants from across the Greater Toronto Area for blood and saliva collection, as well as analysis of saliva via ELISA and sharing of samples (protocol number **23901**). The Unity Health Network (UHN) REB and Mount Sinai Hospital REB granted approval for collection of saliva and blood as well as analysis via ELISA, pseudotyped lentiviral neutralization assays, T-cell and humoral immunity assays (protocol numbers **20-044 UHN, 20-0078-E and 22-0069-E Mount Sinai**). Health Canada and the Public Health Agency of Canada – National Microbiology Laboratory REB approved use of human plasma and serum for Plaque Reduction Neutralization Test (PRNT) under study number **REB 2020-004P**. The Sainte-Justine University Hospital and Research Center (CR-CHUSJ) Research Ethics Board (REB) approved RECOVER protocols under study numbers **MP-21-2021-3035** and **MP-21-2021-3046** which granted approval for recruitment of health care workers (HCWs) from the Greater Montreal area for blood collection, analysis of DNA, RNA, serum and peripheral blood mononuclear cells (PBMCs), as well as sharing serum samples for humoral studies and receiving samples for T cell assays. Serum and saliva samples were obtained from COVID-19 infected individuals confirmed by Polymerase Chain Reaction (PCR) or Rapid Antigen Testing (RAT) by the Toronto Invasive Bacterial Diseases Network (REB studies **20-044 UHN**, **02-0118-U** and **05-0016-C Mount Sinai Hospital**).

### Recruitment and participants – CoVaRR-Net Cohort (Supplemental Table 1)

Healthy adults from the Greater Toronto Area (Ontario, Canada) who had not experienced COVID-19 infection prior to vaccination were recruited and sampled for blood and saliva at the University of Toronto and Unity Health. Sampling was conduced one to three months following either the last dose of vaccine or PCR/RAT confirmed COVID-19 infection. Post-Omicron BT infection subjects (n=16) who had received two vaccine doses (primary series) followed by an Omicron BT infection (confirmed via RAT or PCR) between December 2021 and January 2022 were sampled for saliva, serum, plasma and PBMCs. Three participants were excluded due to negative anti-nucleocapsid antibody levels, resulting in a total of 13 participants. Additional samples collected from healthy adults at Mount Sinai Hospital (resulting in total n=21) were used for serum and salivary analysis of antibody levels only, as no PBMCs were collected for these participants. Samples matched for vaccine type, age and sex were selected after two (primary series, n=33) and three (primary series + booster dose, n=15) doses of vaccine. Participants with insufficient PBMCs sample volume or positive anti-nucleocapsid antibody levels, indicating previous SARS-CoV-2 infection were excluded from this selection.

### Recruitment and participants - RECOVER Cohort (Supplemental Table 2)

HCWs who were infected with the ancestral Wuhan-like strain of SARS-CoV-2 were recruited through five University Hospitals and vaccination centers in the Greater Montreal Area (Quebec, Canada) following their positive PCR result for SARS-CoV-2 infection.^9^ Study participants (n=569) were followed longitudinally for up to two years, and were vaccinated through routine public health programs. Blood samples were collected at an interval of three months. All samples from the RECOVER cohort were processed for serum, plasma and PBMCs isolation and cryopreserved by the Mother-Child Institutional Biobank at the CR-CHUSJ as previously described.^10^ Relevant samples from the RECOVER cohort were selected for this study. Post-Omicron BT infection subjects (n=17) were infected during the same period of infection as the CoVaRR-Net cohort (December 2021 to February 2022), after receiving two doses of vaccine (primary series). Samples matched for sex, age and ethnicity were identified after two (primary series, n=20) and three (primary series + booster dose, n=15) doses of vaccine. Time post-exposure was also matched between the post-BT and post-3^rd^ dose samples. Here, blood samples were collected around six weeks after the third dose of vaccine (booster) or after BT infection. Post-2^nd^ dose samples were collected around 4.4 months after the second dose, at the same period as the other two groups had samples collected. All selected subjects had been vaccinated with the BNT162b2 mRNA vaccine.

### Collection and processing of saliva and blood

Methodology for saliva collection followed our previously published protocol.^6^ Participants were instructed to abstain from eating, drinking, or smoking for at least 30 minutes before providing their saliva samples. Samples were collected using the Salivette system (Sarstedt, Numbrecht, Germany). The Salivette system is comprised of a large tube with a smaller tube that contains a cotton swab. Participants were instructed to chew on the cotton swab for three minutes, after which they returned it to the smaller tube. The entire system was then centrifuged at 1000 x g for five minutes, allowing saliva to filter through a hole in the smaller tube and be collected in the larger tube. The collected saliva was aliquoted into 300 to 500 µl aliquots which were stored at -80°C until ready to use. No viral inactivation methods were taken for these samples, as they were collected from vaccinated, non-infected participants, or after the infectious period in the case of BT infection.

Blood samples were collected using ACD tubes (BD, #364606) for the isolation of PBMCs and SST tubes (BD, #367985) for serum extraction. Plasma was isolated after centrifugation for ten minutes at 1000 x g at room temperature with the brake off. Remaining red blood cells and PBMCs were mixed and diluted 1:1 in PBS. PBMCs were separated from diluted blood by centrifugation over 15 mL of Ficoll (Cytiva, #1714003) in 50 mL modified conical tubes according to manufacturers protocol (STEMCELL, #85450). The PBMCs layer was then decanted into new 50 mL conical and washed by adjusting sample volumes to 50 mL with HBSS (Wisent, #311-512-CL). Cell samples were washed by centrifugation for ten minutes at 300 x g at room temperature. Cell pellets were washed a second time by resuspending and combining replicate cell pellets in a total of 50 mL of HBSS, followed by another centrifugation with the same parameters. Isolated cells were counted and resuspended to approximately 1 × 10^7^ cells/mL in a final freezing solution consisting of 50% v/v fetal bovine serum (Gibco, 12483-020) and 10% DMSO (Sigma-Aldridch, #D2650-100ML) in RPMI (Thermo Scientific, #11875093). Aliquots were stored in liquid nitrogen. SST tubes were centrifuged for ten minutes at 10 000 RPM at room temperature. The isolated serum was then aliquoted into 500 µl aliquots into two mL cryovials (Sarstedt, #72.694.006) and stored at -80°C.

### Enzyme-Linked ImmunoSpot Assay (ELISpot) for the detection of IFN-y secreting T cells

Functional T-cell immunity was measured through Enzyme-Linked ImmunoSpot Assay (ELISpot) for the detection of IFN-γ secreting cells. Frozen PBMCs were rapidly thawed and rested overnight before stimulation. Cells were then stimulated with 1 µg/mL of different SARS-CoV-2 peptides mega pool from the Spike glycoprotein of the ancestral Wuhan-1 like strain or variants B.1.1.7 (Alpha), B.1.351 (Beta), P.1 (Gamma), B.1.617.2 (Delta) and B.1.1.529 (Omicron) from JPT Peptide-Technologies. The assay also included non-spike proteins of SARS-CoV-2 (nucleocapsid and membrane protein) to validate the absence of previous infection in the cohort of presumed uninfected individuals. Spots were revealed and analyzed as previously described by our team.^10^ Culture media and CytoStim^TM^ (AIM-V® Medium (1X), Gibco, #12055-091 and Human CytoStim^TM^, Miltenyi Biotec, #130-092-173) were used as negative and positive controls respectively. The positive threshold response was defined as 25 spot-forming units (SFU) per million of PBMCs.

### Enzyme-linked immunosorbent assays (ELISA) for detecting total IgA and IgG in serum

A chemiluminescent ELISA assay was used to detect antibody levels to full-length spike trimer, its receptor binding domain (RBD) and nucleocapsid as previously described.^21,22^ Briefly, LUMITRAC 600 high-binding white polystyrene 384-well microplates (Greiner Bio-One, #781074 ; VWR, #82051-268) were pre-coated overnight with 10 µL per well of antigen (Ag) : 50 ng spike (SmT1), 20 ng RBD (331-521) and 7 ng nucleocapsid, all supplied by the National Research Council of Canada (NRC). The next day, the assay was performed at room temperature with washing four times in 100 µL PBS-T before each of the following steps. Step one : wells were blocked for 1 h in 80 µL 5% Blocker BLOTTO (ThermoFisher Scientific, #37530). Step two : 10 µL of serum diluted 1:160. 1:2,560 or 1:40,960 (for anti-IgG only) in 1% final Blocker BLOTTO in PBS-T was added and incubated for two hours. Step three : 10 µL of a human anti-IgG fused to HRP (IgG#5 by NRC, 0.9 ng/well) or human anti-IgA conjugated to HRP (Jackson ImmunoResearch, #109-035-127, 0.8 ng/well) diluted in 1% final Blocker BLOTTO in PBS-T was added, followed by a 1-h incubation. Step four : 10 µL of ELISA Pico Chemiluminescent Substrate (ThermoFisher Scientific, #37069, diluted 1:4 in MilliQ distilled H2O) was added and incubated for 5-8 min. Chemiluminescence was measured on an EnVision 2105 Multimode Plate Reader (Perkin Elmer) plate reader at 100 ms/well using an ultra-sensitive detector. Raw chemiluminescent values were normalized to a synthetic standard included on each assay plate (For IgG : VHH72-Fc supplied by NRC for spike/RBD or an anti-nucleocapsid IgG Ab from Genscript, #A02039 ; For IgA : anti-spike CR3022 from Absolute Antibody, #Ab01680-16.0 and anti-nucleocapsid CR3018 from Absolute Antibody, #Ab01690-16.0), and these relative ratios were further converted to binding antibody units (BAU/mL) using the WHO International Standard 20/136 as the calibrant.^22^ Positivity thresholds were determined for the 1:160 dilution using three standard deviation from the mean of control samples as previously described.

### Pseudotyped Lentiviral Neutralization Assays

The generation of spike-pseudotyped lentivirus particles and the pseudovirus neutralization assay was performed as described previously with some modifications.^23^ Pseudotyped lentivirus particles were co-transfected with packaging (psPAX2, Addgene, #12260), the ZsGreen and luciferase reporter construct (pHAGE-CMV-Luc2-IRES-ZsGreen-W, provided by Jesse Bloom), and the Spike protein constructs from the wild-type (Ancestral) bearing the D614G mutation, and the variants of concern (Beta, Delta, Omicron BA.1, BA.2 and BA.5) into HEK293TN cells (System Biosciences, LV900A-1). Viral supernatants were harvested 48 hours post-transfection, and viral titer assay was performed by infecting HEK293T-ACE2/TMPRSS2 cells, followed by a luciferase assay to determine the relative luciferase unit. For neutralization assays, diluted sera samples (1:22.5) were prepared and serially diluted by 3-fold over seven dilutions, followed by incubation with diluted Spike-pseudotyped lentivirus for one hour at 37°C prior to addition to HEK293T-ACE2/TMPRSS2 cells. The infected cells were then incubated for an additional 48 hours, followed by cell lysis and luminescence signal measurement using the Bright-Glo Luciferase Assay System (Promega) and PerkinElmer EnVision 2015 Multimode Plate Reader, respectively, for relative luciferase unit quantifications. Unless otherwise specified, the 50% virus neutralization titer (ID50) values were calculated in GraphPad Prism version 9.5.1 using a nonlinear regression algorithm (log[inhibitor] versus normalized response - variable slope). In patients with an absence of 50% neutralization, a log_10_ ID50 of zero was recorded.

### Plaque reduction neutralization test (PRNT) for testing viral neutralization

Neutralizing antibodies were assessed by PRNT50 and PRNT90 assays as recently described.^24^ In brief, samples were serially diluted and mixed with SARS-CoV-2 wild-type (Ancestral) as well as Omicron BA.1, BA.2 and BA.5 variants. No neutralization, 50% neutralization, 90% neutralization and no virus controls were prepared in a similar fashion. Following a one hour incubation at 37°C, each antibody/virus mixture was added to a culture of Vero E6 cells. Following a three to four days incubation, the cells were fixed and stained with 0.5% crystal violet (w/v) in order to calculate the number of plaques observed in each well. The number calculated was compared to the average plaque count in the 50% and 90% neutralization control wells in order to calculate final PRNT50 and PRNT90 titers for each antibody sample.

### Expression, purification and production of Wuhan, Omicron and SARS-CoV Spike and RBD proteins for detection of anti-Spike and anti-RBD IgA and IgG in saliva

The SARS-CoV-2 Spike and RBD proteins used in these experiments were expressed, purified and biotinylated as previously described.^6,21,25^ The SARS-CoV-1 Spike protein was codon optimized and stabilized by mutating the S1-S2 cleavage site (R667S) and by converting six residues in the S2 region to prolines (F799P, A874P, A881P, S924P, K968P, V969P). The SARS-CoV-2 Omicron variant Spike protein was codon optimized and stablilized by mutating the Furin cleavage site (R682S, R683S, R685S) and by converting six residues in the S2 region to prolines (F817P, A892P, A899P, A942P, K986P, V987P), as described previously.^25,26^ The RBD construct was fused with a 6xHis tag and an AviTag. The soluble ectodomains of the Spike proteins were fused with a foldon trimerization motif, a 6xHis tag and an AviTag. The resulting plasmids were used to transfect FreeStyle 293-F cells for the secretion of the RBD and soluble Spike trimers using previously reported methods.^6,25^ The proteins were purified from the cell culture media using Ni-NTA affinity chromatography. The purified proteins were then site-specifically biotinylated in reactions containing 200 µM biotin, 500 µM ATP, 500 µM MgCl_2_, 30 µg/mL BirA, 0.1% (v/v) protease inhibitor cocktail and not more than 100 µM of the protein-AviTag substrate. The reactions were incubated at 30 °C for two hours and the biotinylated proteins were then purified by size-exclusion chromatography. The qualities of all Spike protein preparations were confirmed by negative stain electron microscopy. The qualities of the RBD preparations were confirmed by the binding to the CR3022 antibody using BioLayer Interferometry.^27^

### Enzyme-linked immunosorbent assays (ELISA) for detecting anti-Spike and anti-RBD IgA and IgG in saliva

An amplified (biotin/streptavidin) ELISA assay was used to detect IgG and IgA antibody levels to full-length Spike trimer and its RBD.^6,21^ Notably we previously showed that salivary anti-Spike and anti-RBD specific salivary IgA highly correlated with anti-Spike/RBD specific salivary SIgA.^6^ Briefly, all plates were coated with 50 µL per well of 2 µg/mL or 20 µg/mL of biotinylated RBD and Spike, respectively. Control wells were coated with sterile PBS. Once coated, the plates were incubated overnight at 4°C, after which the coating solution was discarded, and plates were blocked with 200 µL per well of 5% BLOTTO solution (5% w/vol skim milk powder in sterile PBS) for two hours at 37°C. During this period, samples were thawed and pre-incubated in separate streptavidin-coated plates to account for anti-streptavidin activity in saliva. The blocking solution was then discarded and samples were added at dilutions of 1:5, 1:10 and 1:20 in 2.5% BLOTTO solution for two hours at 37°C to the antigen-coated plates Sample solutions were then discarded and plates were washed three times with 0.05% PBS-Tween (PBS-T, BioShop, #TWN510). This was followed with addition of 50 µL per well of horseradish peroxidase (HRP)-conjugated IgG (Southern Biotech, 2044-05) or IgA (Southern Biotech, 2053-05) at dilutions of 1:1000 and 1:2000 respectively in 2.5% BLOTTO. After adding to the plates, the secondary antibody solution was incubated for one hour at 37°C and, after which the plates were washed three times with 0.05% PBS-T. Lastly, plates were developed by adding 50 µL per well of 3,3’,5,5’ tetramethylbenzidine (TMB) Substrate Solution (ThermoFisher, 00-4021-56) for 90 seconds. Development was stopped by adding 50 µL of 1N H2SO4 to each well. Optical density (OD) was read at a wavelength of 450 using a spectrophotometer (Thermo Multiskan FC).

### Enzyme-linked immunosorbent assays (ELISA) for detecting albumin in saliva

The concentration of albumin in each saliva sample was measured using human albumin detection ELISA kits (Abcam, ab109788). We began by adding saliva samples in three dilutions (1:200, 1:400, 1:800) to a 96-well plate precoated with human albumin, as well as 50 µL per well of the provided albumin standard. The samples were incubated for one hour at 37°C, after which the plates were washed five times using the provided wash buffer. Next, we added 50 µL per well of biotinylated anti-albumin antibody to each well and incubated the plates for 30 minutes at 37°C. The plates were then washed again five times using wash buffer, followed by addition of 50 µL per well of HRP-conjugated streptavidin. The plates were once again incubated for 30 minutes at 37°C and washed five times with wash buffer. Lastly, we developed the plates by adding 50 µL per well of the chromogen substrate for 25 minutes and stopped development using 50 µL per well of stop solution. The OD450 values were read using a spectrophotometer (Thermo Multiskan FC) and a standard curve was used to calculate the concentration of albumin in µg/mL for each sample.

### Enzyme-linked immunosorbent assays (ELISA) for detecting total IgA and IgG in saliva

The concentration of total Ig in saliva was measured as recently described^21^. In brief, 96 well plates were coated with anti-human total Ig and incubated overnight at 4°C. The next day, coating antigen was discarded and plates were blocked using 5% BLOTTO solution for two hours at 37°C. The blocking solution was then discarded and samples and standards were added to the plates in serial dilutions, followed by another two hours incubation at 37°C. Next, the plates were washed using 0.05% PBS-Tween and the detection antibody (either anti-human IgA HRP or anti-human IgG HRP) was added to the corresponding plates and incubated for one hour at 37°C. This was followed by a second wash and development using TMB, followed by 1N H2SO4 to stop the reaction. Optical density was read at a wavelength of OD450 using a spectrophotometer, and standard curves were used to calculate the concentration of total IgA and IgG in µg/mL for each sample.

### Analysis of saliva ELISA Data

The raw OD450 readout for the PBS control well was substracted from the raw antigen specific OD450 value at each dilution (1:5, 1:10, 1:20). These adjusted OD450 values were then used to calculate the area under the curve (AUC) for each sample. This process was also done for the positive controls, which consisted of pooled acute and convalescent COVID-19 patient saliva. We then adjusted each individual sample AUC to a pre-selected standard based on the values calculated for the positive control. The side by side comparison of total IgG, IgA and albumin concentration per sample is reported (**Fig. S3**). Considering that total IgA was variable between the groups while albumin levels were similar, we decided to move forward with normalizing antigen-specific IgA and IgG results to albumin levels.

### Statistics

Statistical tests were performed using Prism 9, version 9.2.0 (2021 GraphPad Software, LLC). Mann-Whitney or Kruskal-Wallis unpaired nonparametric tests were applied to assess statistical significance between groups as variables did not follow a normal distribution. Significance was established as *P<0.05, **P<0.01, ***P<0.001, ****P<0.0001.

## DATA AVAILABILITY

The datasets generated and analysed in the current study are available from the corresponding author upon reasonable request.

## ACKNOWLEDGMENTS

We thank several teams who allowed for this research to be successful, the participants who agreed to participate in these studies, as well as the nurses and research coordinators from each participating centers. We are also very thankfull for the Coronavirus Variants Rapid Response Network (CoVaRR-Net) Biobank and the Sainte-Justine University Hospital Mother Child Biobank personnel for processing and storing the samples from the studies. Special thanks to Xinliu Angel Li, Mohammad Mozafarihashjin, Maxim Lefebvre, Antonio Estacio, Fazia Tadount, Kelsey Adams, Louise Wang and Sylvie Nicholson for their role in participant recruitment and sampling as well as Kim Crasta, Ryan Law, Jocelyne Ayotte, Jessie Beauchemin, Vanessa Truong, Annie Bilodeau and Amal Abdi for their role in sample preparation. We also acknowledge Geneviève Mailhot, Melanie Delgado-Brand, Tulunay Tursun, and Freda Qi for their role in COVID-19 serology analysis. Antigens, protein standards, and secondary antibodies for serum ELISA were kindly provided by The Pandemic Response Challenge Program of the National Research Council of Canada (Dr. Yves Durocher). This collaborative work was initiated and conducted through the Coronavirus Variants Rapid Response Network (CoVaRR-Net). Jennifer Gommerman and Ciriaco Piccirilo lead the Immunology and Vaccine Protection Pillar of CoVaRR-Net. Anne-Claude Gingras leads the Functional Genomics and Structure-Function Pillar of CoVaRR-Net and is the Canada Research Chair (Tier 1) in Functional proteomics. The CoVaRR-Net funds the CoVaRR-Net Biobank that supported this study.

*This study was supported by funding from a CoVaRR-Net Rapid Response Research Grant (MB, CP, CQ, JR, ACG, HD, JG) under CIHR’s Rapid Response Network to SARS-CoV-2 Variants (Funding Reference Number : 175622) as well as the following grants :*

- *Canadian Institutes of Health Research GA1-177703 (ACG), GA2-177710 (JG) and VR2172712 (CQ, HD)*
- *Coalition for Epidemic Preparedness Innovations and Canadian Institutes of Health Research 468231 (HD)*
- *Ontario Together COVID-19 Rapid Response 20013418 (JG)*
- *Public Health Agency of Canada 2122-HQ-000225 (CQ, HD) and 2223-HQ-000261 (JG)*

The robotics equipment used is housed in the Network Biology Collabrative Centre at the Lunenfeld-Tanenbaum Research Institute, a facility supported by the Canada Foundation for Innovation, the Ontario Government, Genome Canada and Ontario Genomics (OGI-139).

## Author contributions

Conceptualization : MB, CP, ACG, HD, JG

Methodology : SN, SSM, AK, QH, HW, KC, ZL, YL BB, WH, JR, ACG, HD, JG

Investigation : ACG, HD, JG

Visualization : SN, SSM, LS, BB, HD, JG

Funding acquisition : HW, MB, CP, CQ, JR, ACG, HD, JG

Project administration : GC, KC, LS, BB

Subject recruitment : AMG, MO, CQ

Supervision : HW, KC, OR, JR, ACG, HD, JG

Writing – original draft : SN, SSM, HD, JG

Writing – review & editing : SN, SSM, QH, HW, KC, LS, ACG, HD, JG

## Conflicts of Interest and Financial Disclosures

ACG has received research funds from a research contract with Providence Therapeutics Holdings, Inc. for other projects.

No other conflict of interest or financial disclosures.

## Notes

### Funding Statement

This study was supported by funding from a CoVaRR-Net Rapid Response Research Grant (MB, CP, CQ, JR, ACG, HD, JG) under CIHR Rapid Response Network to SARS-CoV-2 Variants (Funding Reference Number : FRN 175622) as well as the following grants :
- Canadian Institutes of Health Research GA1-177703 (ACG), GA2-177710 (JG) and VR2172712 (CQ, HD)
- Coalition for Epidemic Preparedness Innovations and Canadian Institutes of Health Research 468231 (HD)
- Ontario Together COVID-19 Rapid Response 20013418 (JG)
- Public Health Agency of Canada 2122-HQ-000225 (CQ, HD) and 2223-HQ-000261 (JG).
The robotics equipment used is housed in the Network Biology Collaborative Centre at the Lunenfeld-Tanenbaum Research Institute, a facility supported by the Canada Foundation for Innovation, the Ontario Government, Genome Canada and Ontario Genomics (OGI-139).

### Author Declarations

The Research Ethics Board of the University of Toronto granted approval for recruitment of participants from across the Greater Toronto Area for blood and saliva collection, as well as analysis of saliva via ELISA and sharing of samples under protocol number 23901. The Reserach Ethics Boards of the Unity Health Network and Mount Sinai Hospital granted approval for collection of saliva and blood as well as analysis via ELISA, pseudotyped lentiviral neutralization assays, T-cell and humoral immunity assays under protocol numbers 20-044 UHN, 20-0078-E and 22-0069-E Mount Sinai. The Research Ethics Board of Health Canada and the Public Health Agency of Canada - National Microbiology Laboratory approved use of human plasma and serum for Plaque Reduction Neutralization Test (PRNT) under study number REB 2020-004P. The Research Ethics Board of the Sainte-Justine University Hospital and Research Center approved RECOVER protocols under study numbers MP-21-2021-3035 and MP-21-2021-3046 which granted approval for recruitment of health care workers (HCWs) from the Greater Montreal area for blood collection, analysis of DNA, RNA, serum and peripheral blood mononuclear cells (PBMCs), as well as sharing serum samples for humoral studies and receiving samples for T cell assays. The Research Ethics Board of the Toronto Invasive Bacterial Diseases Network approved the use of serum and saliva samples from COVID-19 infected individuals under protocol numbers 20-044 UHN, 02-0118-U and 05-0016-C Mount Sinai Hospital.

### Summary of Updates

The manuscript title was revised.

