## Supplementary Material for "Comparison of Omicron Breakthrough Infection Versus Monovalent SARS-CoV-2 Intramuscular Booster Reveals Differences in Mucosal and Systemic Humoral Immunity"

Hélène Decaluwe

CHU Sainte-Justine Research Center

3175, Chemin de la Côte-Sainte-Catherine, Montréal, QC, Canada (H3T 1C5)

Jennifer L. Gommerman

University of Toronto, Medical Sciences Building, 1 King's College Circle, Room 7233, Toronto, ON, Canada (M5S 1A8)

### SUPPLEMENTAL FIGURES AND LEGENDS

#### Supplemental Tables

**Supplemental Table 1.** Characteristics of participants naïve to SARS-CoV-2 prior to vaccination.

**Supplemental Table 2.** Characteristics of participants infected with SARS-CoV-2 prior to vaccination and breakthrough infection.

#### Supplemental Figures

**Figure S1.** Omicron breakthrough infection boosts T-cell responses in both vaccinated only and hybrid immune individuals against all variants assessed.

**Figure S2.** Neutralizing antibody titers against several variants in vaccinated individuals who were previously naïve to the infection assessed through plaque reduction neutralization test.

**Figure S3.** Vaccinated individuals and breakthrough subjects have similar albumin levels in the saliva which allowed for normalization of the response.

**Supplemental Table 1. Characteristics of participants naïve to SARS-CoV-2 prior to vaccination.**

| | Number (%) of individuals with two vaccine doses<br>2x Vax<br>N = 33 | Number (%) of individuals with three vaccine doses<br>3x Vax<br>N = 17 | Number (%) of individuals with two vaccine doses + BT<br>2x Vax + BT<br>N = 21 |
|---|---|---|---|
| **Demographics** | | | |
| **Sex** | | | |
| Male | 11 (33) | 8 (47) | 8 (38) |
| Female | 22 (67) | 9 (53) | 13 (62) |
| **Age (Y)** | | | |
| Mean ± SD | 42 ± 11 | 39 ± 12 | 36 ± 12 |
| Median [Min, Max] | 40 (22, 64) | 35 (25, 59) | 32 (20, 62) |
| **Time since last exposure (vaccine or BT) at analysis (W)** | | | |
| Mean ± SD | 6.2 ± 3.3 | 5.1 ± 2.3 | 5.9 ± 2.0 |
| Median [Min, Max] | 5.4 [1.9, 14] | 4.3 [2,9] | 6.3 [1.6, 8.1] |
| **Vaccination received** | | | |
| Pfizer-BioNTech (BNT162b2) | 8 (24.2) | 4 (23.5) | 11 (52.4) |
| Moderna (mRNA1273) | 6 (18.2) | 2 (11.8) | 3 (14.3) |
| Heterologous mRNA vaccines (BNT162b2 & mRNA1273) | 6 (18.2) | 5 (29.4) | 4 (19.0) |
| mRNA vaccine(s) and AstraZeneca-Oxford (ChAdOx1) | 13 (39.4) | 6 (35.3) | 3 (14.3) |

SD : Standard Deviation

Y : Years

W : Weeks

**Supplemental Table 2. Characteristics of participants infected with SARS-CoV-2 prior to vaccination and breakthrough infection.**

| | Number (%) of individuals with two vaccine doses<br>Inf. + 2x Vax<br>N = 20 | Number (%) of individuals with three vaccine doses<br>Inf. + 3x Vax<br>N = 15 | Number (%) of individuals with two vaccine doses + BT<br>Inf. + 2x Vax + BT<br>N = 17 |
|---|---|---|---|
| **Demographics** | | | |
| **Sex** | | | |
| Male | 5 (25) | 4 (27) | 3 (18) |
| Female | 15 (75) | 11 (73) | 14 (82) |
| **Age (Y)** | | | |
| Mean ± SD | 38 ± 9 | 38 ± 10 | 37 ± 11 |
| Median [Min, Max] | 39 [26, 59] | 36 [26, 59] | 41 [20, 58] |
| **Time since last exposure (vaccine or BT) at analysis (W)** [#] | | | |
| Mean ± SD | 19.0 ± 3.2 | 6.2 ± 2.0 | 6.4 ± 2.2 |
| Median [Min, Max] | 18.6 [13.4, 27.7] | 6.9 [2.4, 9.0] | 5.7 [4.3, 13.3] |
| **Vaccination received** | | | |
| Pfizer-BioNTech (BNT162b2) | 20 (100) | 15 (100) | 17 (100) |

[#] : $p < 0.0001$

SD : Standard Deviation

Y : Years

W : Weeks

**Figure S1. Omicron breakthrough infection boosts T-cell responses in both vaccinated only and hybrid immune individuals against all variants assessed.**

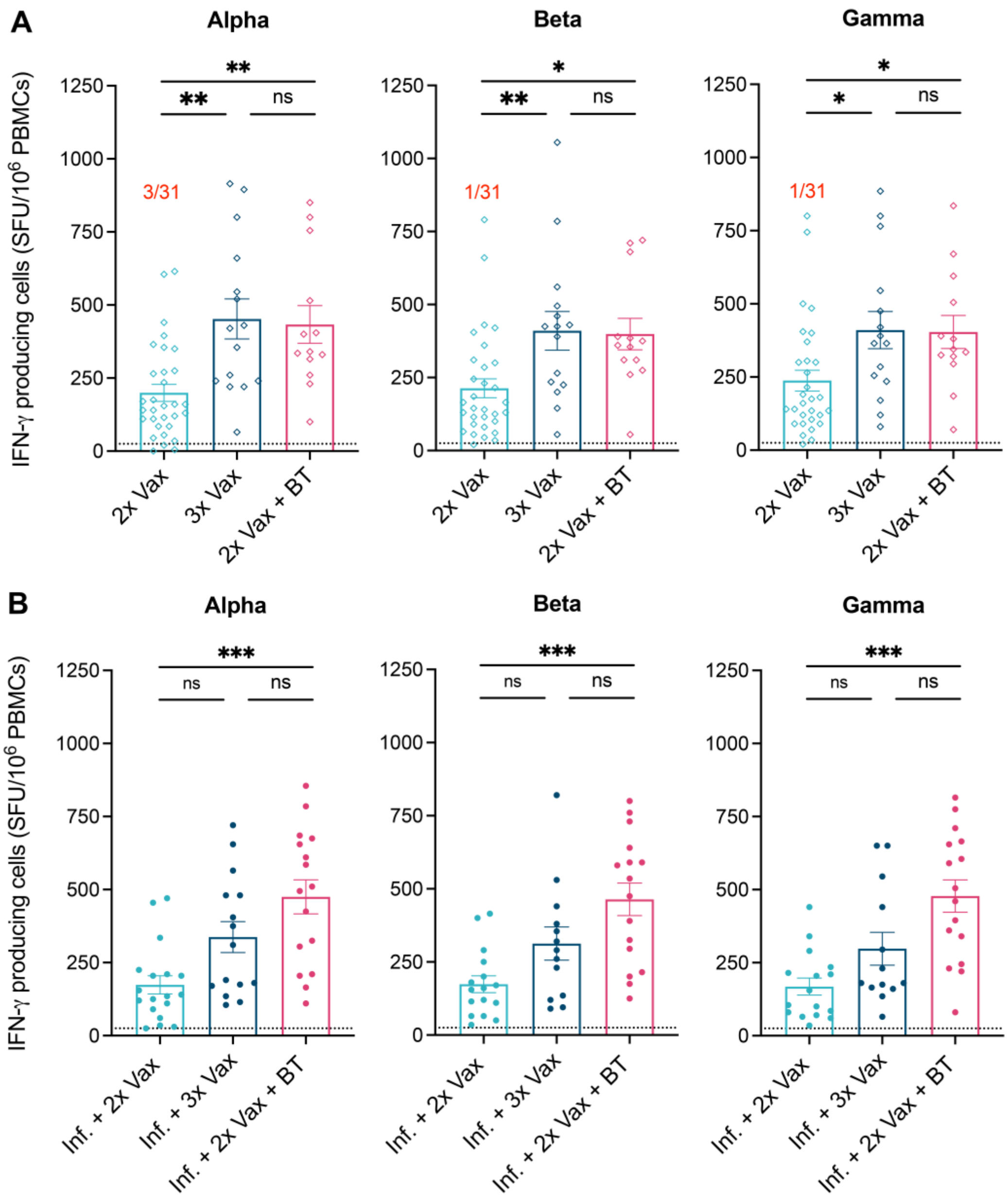


Samples were collected from **(A)** previously naïve individuals (empty diamonds) and **(B)** individuals infected with the ancestral SARS-CoV-2 (full circles). PBMCs were analyzed after two (n=31, light blue), three (n=15, dark blue) or two vaccine doses followed by breakthrough

(BT) infection (n=13, red) for the cohort naïve to SARS-CoV-2. PBMCs were analyzed after two (n=20, light blue), three (n=15, dark blue) or two vaccine doses of BNT162b2 followed by an Omicron BT infection (n=17, red) for the cohort infected (Inf.) prior to vaccination. T-cell responses were assessed by ELISpot after SARS-CoV-2 Spike peptide stimulation with variants strains Alpha (left panels), Beta (middle panels) and Gamma (right panels). Results are expressed in number of IFN-γ producing cells per million PBMCs. Dotted line indicates the positive threshold value. The number of participants with responses under the positive cut-off value are indicated in red for each group when it applies. Error bars indicate mean ± SEM. Statistical significance was established as : ns (not significant) $P > .05$, $^*P < .05$, $^{**}P < .01$, $^{***}P < .001$.

**Figure S2. Neutralizing antibody titers against several variants in vaccinated individuals who were previously naïve to the infection assessed through plaque reduction neutralization test.**

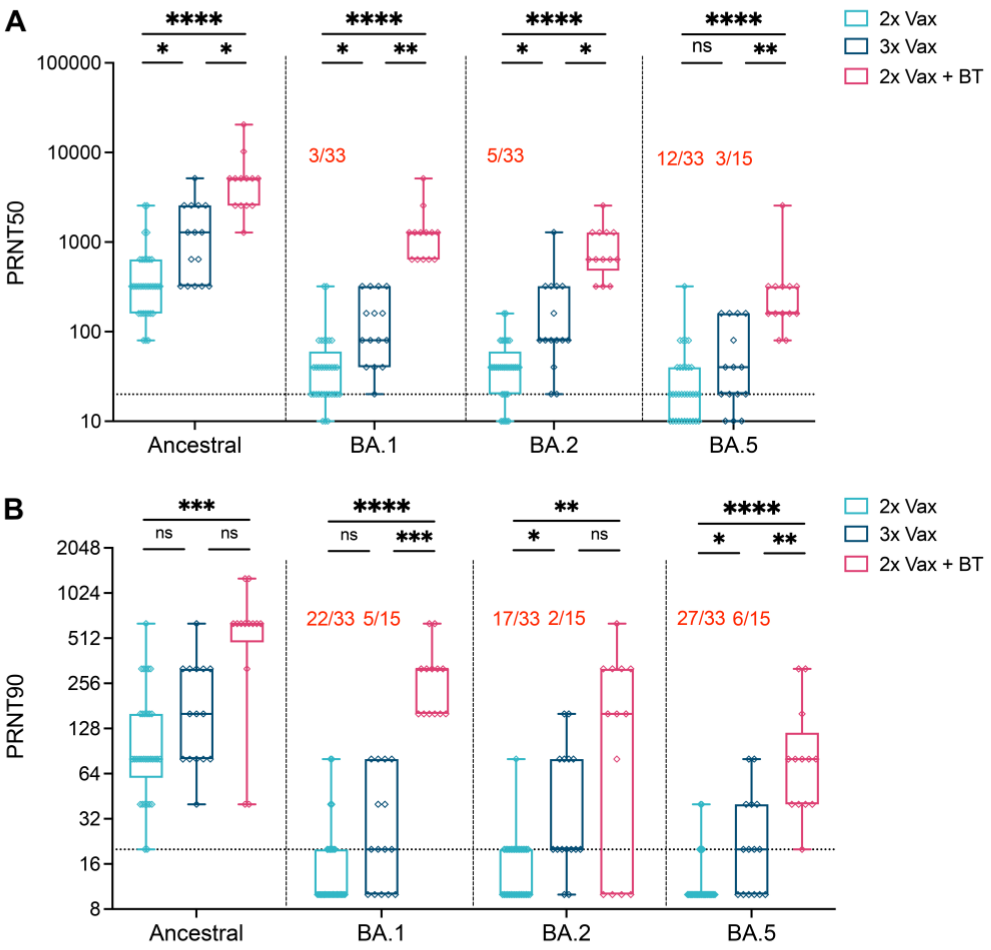


Plaque reduction neutralization tests were conducted to establish the serum dilution that reduced the number of plaques by 50% (**A**) and 90% (**B**). Serums were analyzed after two (n=33, light blue), three (n=15, dark blue) and two vaccine doses followed by an Omicron breakthrough (BT) infection (n=13, red) in individuals who were previously naïve to SARS-CoV-2 (empty diamonds). The number of participants without detectable neutralizing antibodies against specific variants are indicated in red for each group when it applies. Statistical significance was established as : ns (not significant) P >.05, *P <.05, **P <.01, ***P<.001, ****P<.0001.

**Figure S3. Vaccinated individuals and breakthrough subjects have similar albumin levels in the saliva which allowed for normalization of the response.**

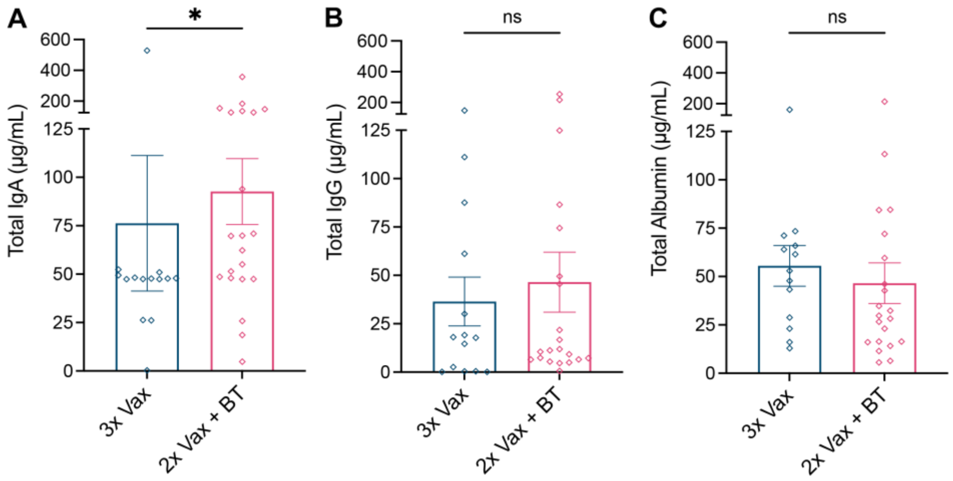


Total IgA (**A**), IgG (**B**) and Albumin (**C**) were measured after three (n=13, dark blue) or two vaccine doses followed by breakthrough (BT) infection (n=21, red) in individuals previously naïve to SARS-CoV-2 (empty diamonds). Error bars indicate mean ± SEM. Statistical significance was established as : ns (not significant) P >.05, *P <.05.
